# MicroRNA-146a polymorphisms are Associated with Psoriasis Vulgaris

**DOI:** 10.1101/2022.09.21.22280226

**Authors:** Kunju Zhu, Shijie Li, Lifan Liang

**Affiliations:** The Department of Biomedical Informatics;School of medicine; University of Pittsburgh, USA; Department of Dermatology, Affiliated Hospital of Guangdong Medical University, Zhanjiang, China

**Keywords:** Psoriasis Vulgaris, MicroRNA-146a, SNPs, Chinese

## Abstract

MicroRNA-146a acts as a critical physiological brake role to prevent the overactivation of inflammatory response pathways and was a key negative regulator of autoimmunity. Ets-1 could bind to the MicroRNA-146a promoter region as a regulator of MicroRNA-146a expression in vitro. IRAK1 is a prominent target of MicroRNA-146a that help it to negatively regulate the release of IL8. This study aimed to investigate the association of MicroRNA-146a, Ets-1(negative regulator) and IRAK1(target) polymorphisms with Psoriasis Vulgaris(***PsV***), the most prevalent chronic inflammatory skin disease in adults, in a Southern Chinese cohort. Seven SNPs in microRNA-146a (rs2431697; rs2910164; rs57095329), Ets-1(rs10893872; rs1128334) and IRAK1(rs1059703; rs3027898) genes were genotyped in 673 subjects (360 PsV cases and 313 controls) by SNaPshot Multiplex Kit (Applied Biosystems Co., USA). We found significant difference in the genotype and allele frequencies of rs2431697 and rs2910164 in MicroRNA-146a gene between the PsV cases and the controls. The dominant model genotype (CC+CT) (*p*=0.019; OR=1.463) and allele (C) (*p*=0.027; OR=1.496) of rs2431697 and the GG genotype (*p*= 0.027; OR=1.582) and allele (G) of rs2910164 were associated with an increased risk of PsV. There was no association of the SNPs with the clinical traits of PsV. Our data provide preliminary evidence that the rs2431697 and rs2910164 polymorphism in the microRNA-146a gene may be involved in the genetic susceptibility to PsV in Southern Chinese. Although further function studies will be required to identify the details of the process, the findings could make a significant step forward in our understanding of the genetic contribution to psoriasis.

**What’s already known about this topic?:**

- MicroRNA-146a is one of the most highly associated MicroRNA to psoriasis.
- MicroRNA-146a acts as a critical physiological brake role to prevent the overactivation of inflammatory response pathways and was a key negative regulator of autoimmunity.

**What does this study add?:**

- Our data provide preliminary evidence that the rs2431697 and rs2910164 polymorphism in the microRNA-146a gene are involved in the genetic susceptibility to PsV in Southern Chinese.

## Introduction

Psoriasis is one of the most common skin disorders resulting from a complex interplay among the susceptibility genes and autoimmune system^1^. Psoriasis vulgaris (PsV), the most common subtype of psoriasis, is characterized by the emergence of hyperkeratosis, parakeratosis and orthokeratosis^2^. However, the molecules pathogenesis of psoriasis is still not completely elucidated.

The microRNAs represent a class of is short and regulatory RNAs that regulate RNA silencing and post-transcriptional process. Recently studies have also shown that the noncoding RNAs (including microRNAs) may be a crucial regulator in human life and so many human diseases^3^. Furthermore, the majority of protein-coding genes may be regulated by microRNAs. Accordingly, microRNAs have been implicated in almost all biological processes, in particular cell differentiation, development and the signaling pathways^3^. In recent years, evidence is accumulating for the role of microRNAs in the pathogenesis of psoriasis rapidly ^4^. Specifically, many studies have confirmed that single nucleotide polymorphisms (SNPs) located in microRNAs genes associated with the susceptibility to psoriasis^4,5^.

Of the susceptibility microRNAs to psoriasis, MicroRNA-146a is one of the most highly associated MicroRNA to psoriasis ^6-8^. Several studies have found that MicroRNA-146a Increased both in the epidermal and dermal compartments of psoriatic skin ^7,9^. Chatzikyriakidou *et al* ^10^ have shown the association between psoriatic arthritis risk in a cohort of patients with psoriasis from Greece and specific polymorphisms found in MicroRNA-146a (rs2910164) and one of its targets IRAK1 (rs3027898, rs1059703). In his study, a very strong association with the rs3027898 IRAK1 variant was also observed ^10^. Specifically, the rs2910164G allele resulted in reduced levels of MicroRNA-146a and impairment of its ability to regulate endothelial growth factor receptor, an important proliferative signal in keratinocytes and psoriasis skin ^11^.

On the other hand, previously study have identified that ETS proto-oncogene 1, transcription factor (ETS1) could affects MicroRNA-146a promoter activity markedly in vitro, by binding the MicroRNA-146a promoter region^12^. Furthermore, knockdown of ETS1 impaired the induction of miR-146a, whereas overexpression of ETS1 enhanced the induction of MicroRNA-146a in vivo experiments. Actually, ETS1 is involved in various immune responses. ETS1 plays a role in immunity, angiogenesis, and cancer progression, and is expressed in B cells, T cells, natural killer cells, endothelial cells, and cancer cells ^13-15^. In T cells, reduction of T helper (Th) 1 cytokine, interferon γ, and decrease in regulatory T cells (Tregs) were observed in Ets1-deficient mice. On the other hand, Th17 differentiation was enhanced by Ets1 deficiency^13-15^. It has also been shown that SNP rs1128334 and rs10893872 located in the 3’-untranslated region (3’-UTR) of Ets-1 are on putative miRNA binding sites and are both associated with SLE in Asian populations^12^. All these results may suggest that Ets-1 influences the activity of MicroRNA-146a and it may take some role in the immunity pathogenesis of psoriasis.

In this study, we aimed to examined the association of MicroRNA-146a, Ets-1(its negative regulator) and IRAK1(its target) gene polymorphisms with PsV in a Southern Han Chinese cohort.

## Materials and methods

### Clinical evaluation and Anthropometric measurements of patients and controls

The detailed method of clinical characteristics evaluation and anthropometric measurements had been described previously^16^. In shortly, General information of all subjects include age, gender, family history, Psoriasis Area and Severity Index (PASI)^17,18^ were recorded. The positive family history was considered if at least one of the first- and second-degree relatives was suffered from psoriasis. PASI score below 6 was defined as mild, and above 6 as severe disease based on median distribution. A total of 360 cases and 313 controls at age of 15 years or more were recruited from 2009 to 2016 in Southern China. The study protocol was approved by the ethical committee of Affiliated Hospital of Guangdong Medical University in compliance with the Helsinki Declaration, and written informed consents were obtained from all participants.

### DNA isolation and genotyping

Genomic DNAs were extracted in accordance with standard protocols from peripheral blood cells by established method^19^. Genotyping of 7 SNPs in microRNA-146a (rs2431697; rs2910164; rs57095329), IRAK1 (rs1059703; rs3027898), Ets-1(rs10893872; rs1128334) genes were determined using SNaPshot Multiplex Kit (Applied Biosystems Co., USA).

### Statistical analyses

*P*<0.05 was the criterion of statistical significance, and all statistical tests were two-sided. Characteristics differences between cases and controls were assessed with the *t* test, the Pearson’s Chi-square test by the SPSS 13.0. The SNP was analyzed for an association with the disease by means of comparison frequency of the minor allele and genotypes, and Hardy–Weinberg equilibrium among controls was confirmed with Chi-squared test for all SNPs. We also performed case-only subtype analyses to examine associations between two significant SNPs genotypes with the clinical traits of PsV.

## Results

### Subjects characteristics

A total of 360 cases (275 men: 185 women; 37.63±15.33 years) and 313 controls (185 men: 128 women; 35.77±14.87 years) were investigated and matched on age and sex (*p*>0.05). For patients, PASI (8.83±7.57), age of onset (30.44±16.06 and 258 Early-onset (≤ 40 years): 102 Later-onset (> 40 years)) and family history (ratio of familial: sporadic cases=75:285) was recorded **(Table 1)**.

**Table 1.** The characteristics of PsV cases and controls.

| **Variables** | **Cases**<br>**(n = 360)** | **Controls**<br>**(n = 313)** | **p-Value** |
| --- | --- | --- | --- |
| Age(mean±sd, years) | 37.63±15.33 | 35.77±14.87 | 0.54 |
| Females, n | 185 | 128 | 0.85 |
| Males, n | 275 | 185 |  |
| PASI score (mean±sd) | 8.63±7.57 | - | - |
| Mean age of onset<br>(mean±sd, years) | 30.44±16.06 | - | - |
| Early-onset (≤ 40 years) , n | 256 | - | - |
| Later-onset (> 40 years), n | 99 | - | - |
| Familial cases, n | 74 | - | - |
| Sporadic cases, n | 281 | - | - |

### Association between seven polymorphisms and the risk of PsV

The genotype frequency distributions for 7 SNPs investigated were consistent with Hardy-Weinberg equilibrium in the control group (*p*>0.05). The comparison of allele distributions revealed that two SNPs (rs2431697, rs2910164) of them were significant associated with the risk of PsV (*p*<0.05). For rs2431697, the minor allele C was present in 15.56% of patients and 11.18% of controls; and significant differences was also observed in dominance model (*p*<0.01), but the recessive model is not. Then for rs2910164, the minor allele G was present in 43.75% of patients and 36.90% of controls; and significant differences was also observed in recessive model (*p*<0.01), but the dominance model is not (**Table 2)**.

**Table 2.**
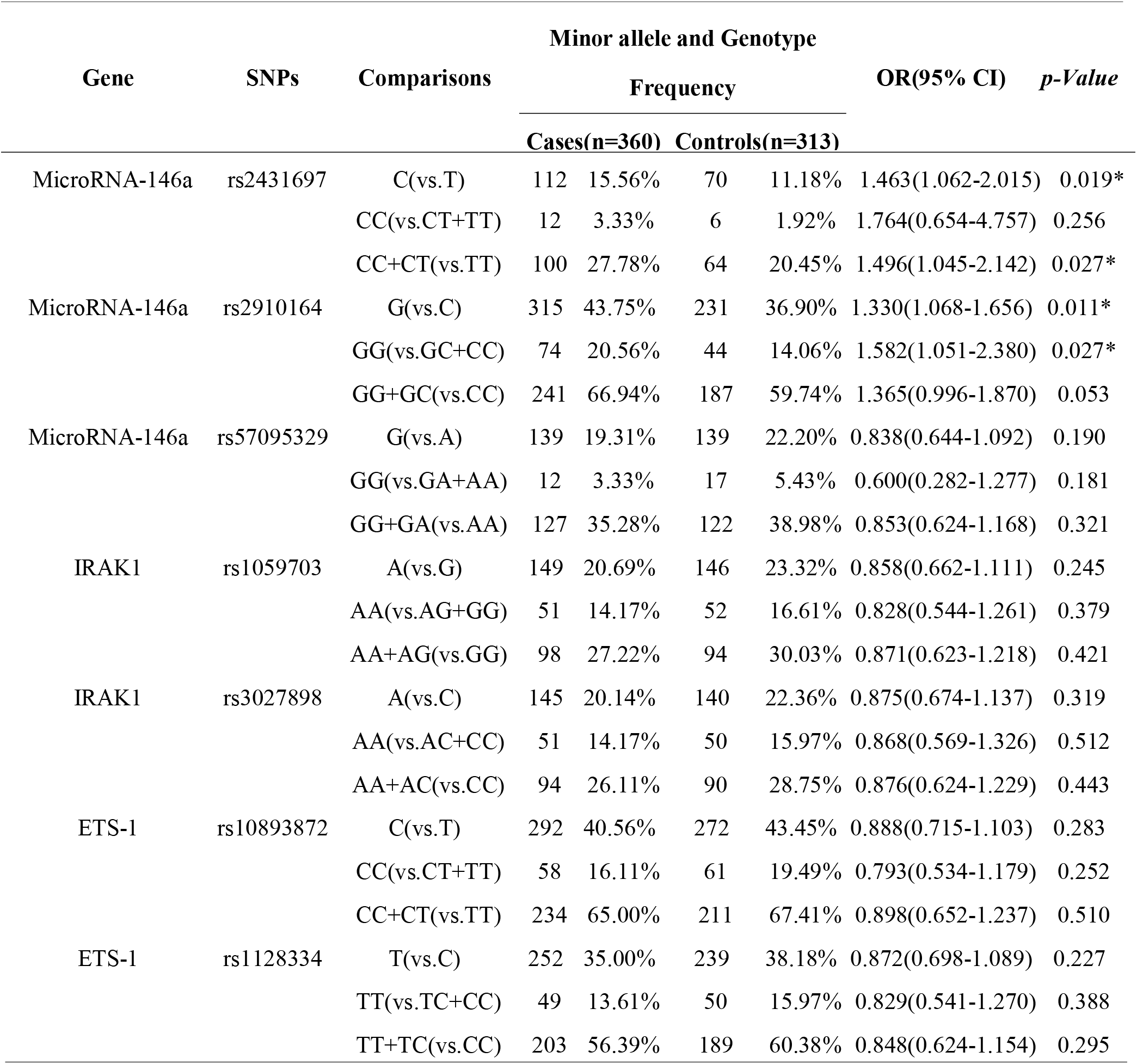
OR estimates of effects on the risk of PsV.

| Gene | SNPs | Comparisons | Minor allele and Genotype |  |  |  | OR(95% CI) | *p*-Value |
| --- | --- | --- | --- | --- | --- | --- | --- | --- |
|  |  |  | Frequency |  |  |  |  |  |
|  |  |  | Cases(n=360) |  | Controls(n=313) |  |  |  |
| MicroRNA-146a | rs2431697 | C(vs.T) | 112 | 15.56% | 70 | 11.18% | 1.463(1.062-2.015) | 0.019* |
|  |  | CC(vs.CT+TT) | 12 | 3.33% | 6 | 1.92% | 1.764(0.654-4.757) | 0.256 |
|  |  | CC+CT(vs.TT) | 100 | 27.78% | 64 | 20.45% | 1.496(1.045-2.142) | 0.027* |
| MicroRNA-146a | rs2910164 | G(vs.C) | 315 | 43.75% | 231 | 36.90% | 1.330(1.068-1.656) | 0.011* |
|  |  | GG(vs.GC+CC) | 74 | 20.56% | 44 | 14.06% | 1.582(1.051-2.380) | 0.027* |
|  |  | GG+GC(vs.CC) | 241 | 66.94% | 187 | 59.74% | 1.365(0.996-1.870) | 0.053 |
| MicroRNA-146a | rs57095329 | G(vs.A) | 139 | 19.31% | 139 | 22.20% | 0.838(0.644-1.092) | 0.190 |
|  |  | GG(vs.GA+AA) | 12 | 3.33% | 17 | 5.43% | 0.600(0.282-1.277) | 0.181 |
|  |  | GG+GA(vs.AA) | 127 | 35.28% | 122 | 38.98% | 0.853(0.624-1.168) | 0.321 |
| IRAK1 | rs1059703 | A(vs.G) | 149 | 20.69% | 146 | 23.32% | 0.858(0.662-1.111) | 0.245 |
|  |  | AA(vs.AG+GG) | 51 | 14.17% | 52 | 16.61% | 0.828(0.544-1.261) | 0.379 |
|  |  | AA+AG(vs.GG) | 98 | 27.22% | 94 | 30.03% | 0.871(0.623-1.218) | 0.421 |
| IRAK1 | rs3027898 | A(vs.C) | 145 | 20.14% | 140 | 22.36% | 0.875(0.674-1.137) | 0.319 |
|  |  | AA(vs.AC+CC) | 51 | 14.17% | 50 | 15.97% | 0.868(0.569-1.326) | 0.512 |
|  |  | AA+AC(vs.CC) | 94 | 26.11% | 90 | 28.75% | 0.876(0.624-1.229) | 0.443 |
| ETS-1 | rs10893872 | C(vs.T) | 292 | 40.56% | 272 | 43.45% | 0.888(0.715-1.103) | 0.283 |
|  |  | CC(vs.CT+TT) | 58 | 16.11% | 61 | 19.49% | 0.793(0.534-1.179) | 0.252 |
|  |  | CC+CT(vs.TT) | 234 | 65.00% | 211 | 67.41% | 0.898(0.652-1.237) | 0.510 |
| ETS-1 | rs1128334 | T(vs.C) | 252 | 35.00% | 239 | 38.18% | 0.872(0.698-1.089) | 0.227 |
|  |  | TT(vs.TC+CC) | 49 | 13.61% | 50 | 15.97% | 0.829(0.541-1.270) | 0.388 |
|  |  | TT+TC(vs.CC) | 203 | 56.39% | 189 | 60.38% | 0.848(0.624-1.154) | 0.295 |

### Association between two risk SNPs (rs2431697, rs2910164) associated with the clinical

We further performed a stratified analysis according to the age of onset, family history and PASI subphenotypes to detect the distribution of the rs2431697 and rs2910164 genotypes in clinical phenotype of PsV. However, there were no significant differences found in subphenotypes of family history or PASI between patients positive and those negative for a particular phenotype **(Table 3)**.

**Table 3.** OR estimates of risk SNPs genotypes on the clinical traits of PsV.

| SNPs | Genotypes<br>Frequency<br>of Cases | Early-onset ($\leq 40$ years) VS.<br>Later-onset (> 40 years) | | Familial cases VS.<br>Sporadic cases | | Mild (PASI $\leq 6$) VS.<br>Severe(PASI >6) | |
| --- | --- | --- | --- | --- | --- | --- | --- |
|  |  | *p* value | OR(95% CI) | *p* value | OR(95% CI) | *p* value | OR(95% CI) |
| rs2431697 |  |  |  |  |  |  |  |
| CC | 12(3.33%) |  | 1 (referent) |  | 1 (referent) |  | 1 (referent) |
| CT | 88(24.44%) | 0.261 | 0.495(0.143-1.716) | 0.251 | 2.25(0.603-8.396) | 0.693 | 1.278(0.377-4.336) |
| TT | 260(72.22%) | 0.325 | 0.557(0.171-1.811) | 0.298 | 1.864(0.541-6.418) | 0.520 | 1.466(0.454-4.739) |
| rs2910164 |  |  |  |  |  |  |  |
| GG | 74(20.56%) |  | 1 (referent) |  | 1 (referent) |  | 1 (referent) |
| CG | 167(46.39%) | 0.056 | 1.880(0.978-3.611) | 0.338 | 0.704(0.343-1.447) | 0.562 | 0.850(0.492-1.471) |
| CC | 119(33.06%) | 0.244 | 1.509(0.754-3.023) | 0.276 | 0.659(0.311-1.400) | 0.830 | 1.066(0.596-1.905) |

OR: odds ratio;
CI: confidence interval;
PASI: Psoriasis Area and Severity Index.
Based on median distribution, PASI $< 6.0$ was defined as mild psoriasis and 6.0 as severe psoriasis.

## Discussion

We conducted the study to evaluate the association between 7 SNPs of MicroRNA-146a gene, Ets-1(its negative regulator) and IRAK1(its target) polymorphisms and risk factors on PsV.

As we know, MicroRNAs are an abundant class of small noncoding RNA molecules, and have been recently recognized as an important regulator in immune homeostasis ^3^. As MicroRNA-146a is a crucial negative regulator of the immune response by negatively regulating *NF-*κ*B* dependent inflammatory signals via direct targeting of IL-1 receptoreassociated kinase 1 (IRAK1) ^20,21^. *NF-*κ*B* is a protein transcription factor and a key regulatory element in a variety of immune and inflammatory pathways, in cellular proliferation and differentiation. Furthermore, *NF-*κ*B* has been hypothesized to connect the altered keratinocyte and immune cell behavior that characterizes the psoriatic milieu ^21,22^.

Moreover, recent studies have demonstrated that SNPs located either in the pre-miRNAs or within miRNA binding sites may contribute to the susceptibility to diseases. Within MicroRNA-146a, several SNPs (rs2910164 and rs2431697) which could lead to the alteration of microRNA function or expression have been noted^23,24^. Specifically, rs2910164 in the pre-miRNA sequence is thought to result in reduced levels of both the pre and mature miRNA. And previously studied found the association between psoriatic arthritis risk in a cohort of patients with psoriasis from Greece and specific polymorphisms found in microRNA-146a (rs2910164) and one of its known targets, IRAK1 (rs3027898, rs1059703) ^10^. In contrast, a separate study showed that the rs2910164 microRNA-146a allele was associated with increased psoriasis susceptibility in Han Chinese patients^11^. The rs2910164 variant C-allele also play a role in the progression of PsA in the South African Indian population^25^. Specifically, the rs2910164G allele resulted in reduced levels of microRNA-146a and impairment of its ability to regulate endothelial growth factor receptor, an important proliferative signal in keratinocytes and psoriasis skin^11^. Srivastava *et al* report protective association of a functional polymorphism in the MicroRNA-146a precursor (rs2910164). Genetic deficiency in MicroRNA-146a leads to earlier onset and exacerbated pathology of skin inflammation, with increased expression of IL-17–induced keratinocyte-derived inflammatory mediators, epidermal hyperproliferation, and increased neutrophil. infiltration. Moreover, they also shown that MicroRNA-146a –deficient mice do not resolve inflammation after discontinuation of imiquimod challenge. The overexpression of MicroRNA-146a suppressed, whereas its inhibition enhanced, IL-17–driven inflammation in keratinocytes. Functionally, MicroRNA-146a impairs the neutrophil chemoattractant capacity of keratinocytes. Finally, delivery of MicroRNA-146a mimics into the skin leads to amelioration of psoriasiform skin inflammation, decreased epidermal proliferation, and neutrophil infiltration. Their results define a crucial role for MicroRNA-146a in modulating IL-17–driven inflammation in the skin^23^.

In addition, strong evidence indicates that rs2910164 leads to increased or decreased microRNA-146a expression in a cell type-dependent manner. For instance, Jazdzewski et al. reported that rs2910164 C allele displayed a 1.8-fold lower level of mature MicroRNA-146a compared to those harboring the G allele^26^. Likewise, Xu et al. found that the G-allelic MicroRNA-146a precursor yielded more mature MicroRNA-146a compared to the C allelic precursor in the 293T cell line^27^. On the other hand, Shen et al. found that MicroRNA-146a expression was 60% higher in the C allele than the G allele in the MCF-7 cell, a model system for breast cancer ^28^. The disparate effects of the rs2910164 C allele on the expression level of MicroRNA-146a may explain the equally disparate effects the SNP has on autoimmune diseases that are localized to different tissues or affect different cell types such as psoriasis and ankylosing spondylitis^24^.

In our study, we found the allele and genotypes of rs2910164 and rs2431697 of MicroRNA-146a associated with the risk of PsV. The associations of the other SNPs within the susceptibility loci remain unclear and further studies are necessary to achieve better understanding the genetic components of PsV. Despite the fact that the function experiments were not carrying out to further illustrate this phenomenon from the level of genetic epidemiology. The findings could make a significant step forward in our understanding of the genetic contribution to psoriasis.

## Data Availability

All data produced in the present study are available upon reasonable request to the authors

## Abbreviations

SNPs: single nucleotide polymorphisms
PsV: psoriasis vulgaris
PBMCs: peripheral blood mononuclear cells
NF-κB: nuclear factor-kappaB
IRAK1: IL-1 receptor associated kinase 1
TNF-α: tumor necrosis factor-α
IL: interleukin
CI: confidence internal
PASI: Psoriasis Area and Severity Index.

## Acknowledgment

We are most grateful to the members of all the patients, the controls and their families who have so willingly participated in this study. This work was supported by grants from the China Natural Science Foundation **(No. 81301351)**. The funders had no role in study design, data collection and analysis, decision to publish, or preparation of the manuscript.

